# Ventilation and the SARS-CoV-2 Coronavirus *Analysis of outbreaks in a restaurant and on a bus in China, and at a Call Center in South Korea*

**DOI:** 10.1101/2020.09.11.20192997

**Authors:** Björn Birnir

## Abstract

In a previous paper [10] a model of the distribution of respiratory droplets and aerosols by Lagrangian turbulent air-flow was developed. It is used to show how the SARS-CoV-2 Coronavirus can be spread by the breathing of single infected person. The model shows that the concentration of viruses in the cloud, exhaled by one person, can increase to infectious levels within a certain amount of time, in a confined space where the air re-circulates. In [10] the model was used to analyze the air-flow and SARS-CoV-2 Coronavirus build-up in a restaurant in Guangzhou, China [23, 22]. In this paper, we add the analysis of two more cases, an outbreak among lay-Buddhists, on a bus [30], traveling to a ceremony in Zhejiang province, China, and an outbreak in a Call Center in Seoul, Korea [24]. The analysis and comparison of these three cases, leads to the conclusion that the SARS-CoV-2 Coronavirus attacks in two steps: The first step is a linear spread between individuals with a couple of days delay. The second step is an polynomial spread effected by the air-conditioning system affecting a much larger number of people. Thus in the second step, the ventilation can become the super-spreader.

## 1 Introduction

The understanding of the transmission of the SARS-CoV-2 Coronavirus has evolved considerably during the current pandemic. Initially, it was thought that virus was mainly spread by infected people coughing or speaking, by large drops that quickly fell to the ground. This understanding lead to the 2 meter (6 feet) physical distancing rule that has been used effectively all around the world to limit the transmission of the disease. It was then shown that the droplets could travel by a turbulent cloud when a person forcefully coughs and that this cloud could travel up to a distance of 7 to 8 meters (25 feet) [11, 29, 12]. It was also argued that smaller droplets and aerosols could be infectious, [4] and the virus was found in small droplets and aerosols in the ventilation system in hospitals in Wuhan [18] and in the air in hospital rooms where Covid-19 patients were being treated [26, 27]. It has also been understood for some time now that the boundary between droplets and aerosols is not sharp, but the droplets and aerosols form a continuum, and evaporation can decrease the size of larger droplets and turn them into airborne droplets and aerosols. This process is of course aided by the ventilations system, in particular when it is heating the air.

The discussion of small droplets and aerosols has mostly been been centered on coughing and sneezing, and speaking, and it has not been sufficiently emphasized that small droplets and aerosols are spread by the breathing of an infected person and their concentration can increase over time, in a confined space. In [10] we developed a model, based on Lagrangian fluid dynamics, of how the cloud of small droplets and aerosols spreads after being exhaled by an infected person and how the concentration of the droplets and aerosols containing the SARS-CoV-2 Coronavirus, can increase over the time the infected person spends in a confined space. The method was used in [10] to analyze an outbreak in a restaurant in Guangzhou, China, on Chinese New Year in January 2020, see [23] and [22]. In this paper we use the method to analyze two more cases, an outbreak among lay-Buddhist traveling to ceremony in Ningbo city, Zhejiang province, in China, in January 2020, see [30], and an outbreak in a Call Center in Seoul, Korea, at the end of February beginning of March 2020, see [24]. These three cases have become examples of outbreaks where the physical distancing rules do not seem to suffice to prevent the infection of a large number of people, see https://english.elpais.com/spanish_news/2020-06-17/an-analysis-of-three-covid-19-outbreaks-how-they-happened-and-how-they-can-be-avoided.html.

It is currently recognized the aerosols play a role in the infection by the SARS-CoV-2 Coronavirus, see the scientific brief from the US Center for Disease Control (CDC) [16], from October 2020. It warns about three possible situations for airborne infection:

- Enclosed spaces within which an infectious person either exposed susceptible people at the same time or to which susceptible people were exposed shortly after the infectious person had left the space.
- Prolonged exposure to respiratory particles, often generated with expiratory exertion (e.g., shouting, singing, exercising) that increased the concentration of suspended respiratory droplets in the air space.
- Inadequate ventilation or air handling that allowed a build-up of suspended small respiratory droplets and particles.

In this paper we quantify the build-up of the aerosols (droplets and particles) in the three case studies mentioned above and show how it can be avoided. It turns out that in most case the small droplets quickly evaporate into aerosols. It is possible that the waves of new infections seen in the Southwest of the United States in late summer, in the Mid-West in the early fall and along the North East Coast as well as along the West Coast in late fall, and California in December, are due to the first hot and then cold weather. This weather forced people to move inside and to spend more time in confined spaces, where the aerosols can build-up, if ventilation is lacking. Low relative humidity, aided by heating, also helps to make these spaces more infectious. To understand this and to improve the ventilation may be key to prevent further surges, before vaccination can alleviate the pressure of new infections.

The paper is organized in the following way: In Section 2 we briefly recall the analysis of the restaurant in Guangzhou, China, in [10], in Section 3 we apply the method to the buses in Ningbo city, China, and in Section 4, to the Call Center in Seoul, Korea. Section 5 contains our discussion and conclusions. In the Appendix A, we explain the more technical details of the method.

## 2 The Restaurant in Guangzhou

On January 24th four members of a family, that had travelled the previous day from Wuhan, ate lunch at a restaurant, in Guangzhou, China. Two other families were present, see Figure 1, in the same part of the restaurant, see [23]. The first family contained one person (A1) who fell ill later the same day and went to the hospital. 12 days later 9 other members of the three families had fallen ill with Covid-19. The infection of these people is consistent with droplet or aerosol transmission because no one else at the restaurant nor the servants fell ill. Only the persons in the direct airstream of the air-conditioner fell ill, most in spite of the fact that the distance between them and the infected person was greater than 2 meters.

**Figure 1:**
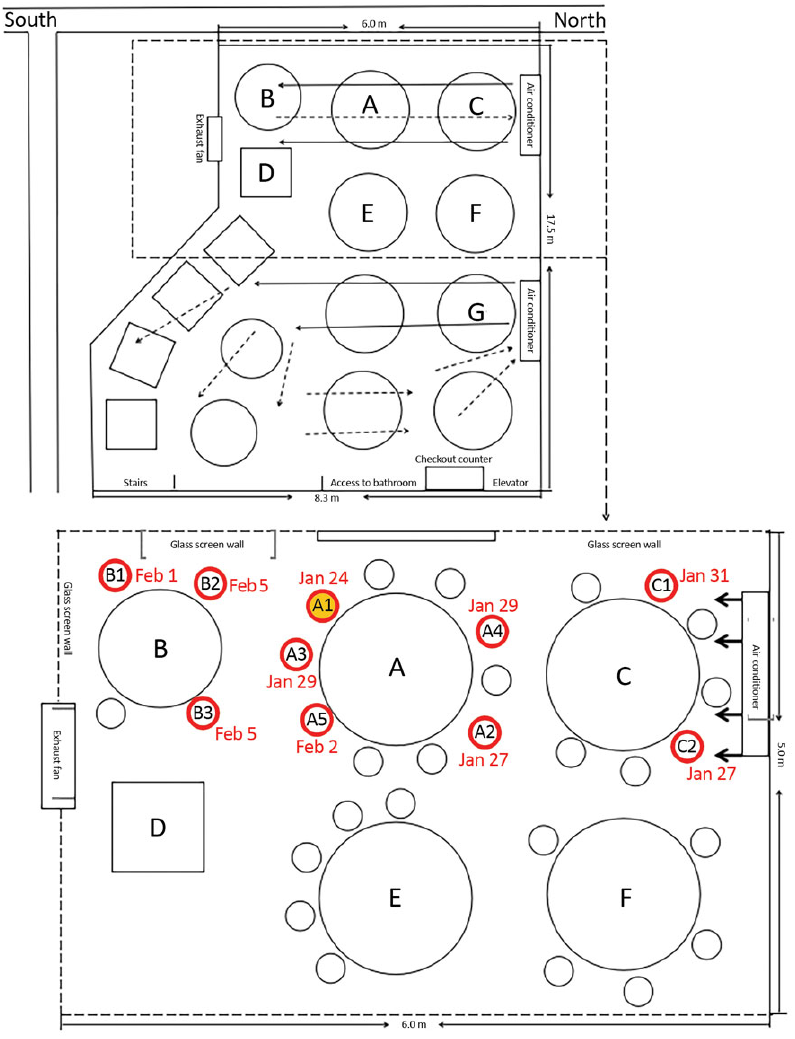
A sketch showing the arrangement of restaurant tables and air conditioning airflow at the site of an outbreak of 2019 coronavirus, in Guangzhou, China, 2020. Red circles indicate where patients that developed the disease were located; yellow-filled red circle (A1) indicates the location of the infected patient. The figure is taken from [23].

In [10] we developed a method to analyze the concentration of the droplets and aerosols containing the SARS-CoV-2 Coronavirus. The method is based on the observation that the small droplets and aerosols can be considered to be passive scalars, or particles that are carried by the flow but do not influence it, in the turbulent Lagrangian airflow in the restaurant, see [10].

The first step in the analysis is to figure out the volume of air in the restaurant. This we get by simply using the dimensions of the space were the three families sat. Then we must compute the volume of the SARS-CoV-2 Coronavirus cloud that the infected person exhales over time and divide the volume of the cloud by the total volume of the air in the restaurant. This gives the concentration of the small droplets or aerosols that every person in the restaurant is exposed to, assuming that we have normalized the concentration to be one, initially, at the infected person. This assumes that if a person is exposed to a concentration one, see [10], over a sufficiently long time interval, if suffices for that person to become infected, as experience has shown.

It makes sense that if an infected person is in a confined space and continues to exhale a cloud of droplets and aerosols, over time, the concentration of the virus-laden drops and aerosols will increase. But how much depends on how often the infected person breathes and what the dimensions of the exhaled cloud are. To determine this one has to use the (Lagrangian) velocity of the air and the dimension of the room to determine two fundamental quantities: These are the Taylor-Reynolds number *Re*_λ_ and the energy dissipation ε of the turbulent cloud. These will in turn determine the time-scale and the rate at which the cloud is expanding and propagating.

We will now summarize the analysis of the restaurant in [10]. The volume of the section of the restaurant, where the infected persons sat, is 3 by 6 by 3.14 meters = 56.52 *m*^3^, see [22]. The Lagrangian air velocity is 0.25 because the air-conditioners are set so that the restaurant guests experience this wind velocity, see [1]. The system size is 3 meters: the distance from the infected person to the wall where the air-conditioner is mounted, see Figure 1. This makes the Taylor-Reynolds number *Re*_λ_ = 705, assuming the temperature is around 20 degrees C^*o*^. The energy dissipation ε = 1.2, is taken from the experimental and simulation data in [5], it gives the Kolmogorov time scale τ_η_ = 3.55 ms (milliseconds). This means that the turbulent cloud containing the SARS-CoV-2 Coronavirus is evolving very fast. The infected person exhales approximately 12 times a minute and each exhale event last 1.2 seconds, or 1/4 of the breathing cycle that lasts 5 seconds. This is the most extreme exhale event, where the infected person coughs and the cough lasts 1.2 seconds, see [14]. It creates the greatest concentration of droplet/aerosols that we will normalize to one. Multiplying this time period with the Lagrangian velocity 0.25 m/s, we get the spacial length of the exhaled cloud, 0.3 m. Computing the area under the curves (blue and red) in Figure 3 (left), and rotating it around the *x* axis, we get the volume, 0.132 m^3^. The cloud has a vase shape, see Figure 3 (left), but its volume is equivalent to the volume of a cylinder of radius 0.264 meters and length 0.6 meters, see Figure 5. If we let the exhaled clouds be contained in a cylinder with this radius extending the length of the contamination area we get the volume 1.32 *m*^3^ for this cylinder. Namely, the length of the cylinder, 6 meters, is 10 times the length of the exhaled cloud, 0.6 meters. The droplet/aerosols in the cylinder now get spread to the pyramid, by the air-conditioning and heat conduction, see Figure 5. This decreases their concentration by a factor of 18.48*/*1.32 = 14. However, the infected person keeps exhaling a new droplet/aerosol cloud every 5 seconds and since the extent of these clouds is 0.6 meters he needs to exhale 10 clouds to fill the 6 meter long cylinder. This takes 50 seconds or 0.83 minutes. In one hour he has filled the cylinder 72 = 60/0.83 times. Thus with no droplet/aerosols lost, the exhaust fan was closed see [22], the resulting concentration in the pyramid is 72 × 1*/*14 = 5.16 times what it was in the cylinder, the first time it was full. In other words, the concentration for everyone sitting at the three tables A, B and C, in Figure 1, in less than 12 minutes, is what it would be if they were sitting next to the infected person. In one hour, the concentration is more than five times what it was initially, if every person on the three table was sitting next to the infected person.

The linear growth of the concentration from the Lagrangian simulation in Figure 3 (left), is an upper bound for the growth of the aerosol concentration as explained in Appendix A. We must solve the ordinary differential equation (ODE) (A.3), where the parameter exhalation rate per volume is given by the Lagrangian simulation. The decay rate of the virions dampens the concentration. The solution of the ODE is compared with the Lagrangian upper-bound in Figure 6, see also Equation 2.1, below.

The vents in the restaurant were closed so there was no ventilation and we can take ventilation coefficient *k*_*v*_ = 0. The droplets settle over time, but they were being blown around by the air conditioner and quickly evaporate into aerosols. This process was presumably helped by modest heating. The estimates of the settling time range from 8-14 minutes, see [32] and [31], whereas the evaporation of the droplets only takes a few seconds, see [19] and [32]. Thus the settling time is just relevant for the process of turning droplets into aerosols. The settling of the aerosols is negligible, even if they settle on the tables they are being blown around by the airflow, and by restricting *C* be the concentration of aerosols, including the nuclei of the evaporated droplets, we can take the settling coefficient *k*_*s*_ = 0. This leaves the decay time of the virions in aerosols. This time is estimated experimentally in [33], [15] and [13]. It depends on the relative humidity and is similar to that of Influenza A. Since the dependence of the latter on the relative humidity is known, we will use the decay coefficients for Influenza A [34] as a proxy. The relative humidity in Guangzhou in January is 72% but the weather is cold and the relative humidity in a heated restaurant can be expected to be much less. We pick it to be 21% and use the corresponding value of the virion decay coefficient *k*_*d*_ = 0.0031 from [34]. Higher values will give qualitatively similar theory, but slightly different quantitatively, see Appendix D in [10]. The aerosol concentrations now lies between the two polynomials

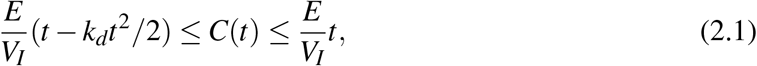

where 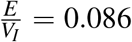 is the exhalation rate in minutes, divided by the volume of the pyramid, see Figure 5. It continues to increase through the time interval and reaches its maximum *C*(60) = 4.72 at the end of the hour, see Figure 6.

If we use the 65% relative humidity in [33] and their corresponding measured value of *k*_*d*_ = 0.01, the picture does not change much, *C*(*t*) continues to increase through the hour and reaches its maximum *C*(60) = 3.87 at the end of it, see Appendix D, in [10].

This quadratic build-up of the concentration of the droplet/aerosols containing the SARS-CoV-2 Coronavirus, in Equation (2.1), shown in Figure 6, is the likely reason for the outbreak in the restaurant in Guangzhou. It shows that in one hour, the rear of the restaurant has become very infectious. Consequently, almost half of the people occupying this space became infected. It they had stayed one more hour, most likely everyone in the space, for that length of time, would have been infected.

It is also shown in [10] that fixing the air-conditioning in the restaurant does not solve the problem. If the recommended amount of fresh air, which is 8 liters per second, per person, is added, the time until the concentration is one everywhere in the contaminated section of the restaurant increases to two hours. After one hour the concentration is 0.5 times what it would be initially sitting next to the infected person. This is still infectious. Moreover, in [10], it is shown that more than 5 times the recommended amount of air has to be injected, into the section of the restaurant where the three families were sitting, for the concentration of the small droplets and aerosols not to build up to infectious levels.

## 3 The Bus with the Lay-Buddhists

On January 19, 2020, 128 persons, lay-Buddhists and 2 bus drivers, took two buses from a neighborhood in Ningbo city, Zhejiang province, in China, to a temple in Ningbo city, 50 min. each way, for a total round trip of 100 min., see [30]. They attended a 150 min. ceremony at the temple that included an indoor lunch, in a spacious room, that lasted 15-30 min. In addition, there were 172 lay-Buddhists that traveled themselves (did not take the two buses) to the ceremony, and participated in it, along with five Buddhist monks for a total of 300 persons.

Of the two buses, Bus 1 contained 60 people, none of whom got sick, Bus 2 contained 68 people, including the infected person a female lay-Buddhist that had shared a meal the previous night with 4 people that had traveled to Hubei province (where Wuhan is located). She was asymptomatic during the trip to the temple but began to feel symptoms that night. Three days later her whole family had become sick and went to the hospital. On the day of the trip, January 19th, there had been no reported cases of the coronavirus in Ningbo city.

Whereas no one on Bus 1 got infected with the coronavirus, on Bus 2, 24 individuals got infected. Of the 172 additional lay-Buddhists attending the event 7, all of whom reported close contact with infected person, got infected. None of the monks got infected. During the event, the passengers in Bus 1 (in fact, both buses) were randomly dispersed among the other participants in the ceremony.

The analysis of the airflow in the bus is analogous to the analysis of the airflow in the restaurant in [10], and summarized above. The air-conditioning on both buses was in re-circulation mode, so no outside air was taken in the through the air-conditioning. The air outside was a chilly 0-10 *C*^*o*^, so the air-conditioning was heating and the air rose towards the ceiling of the buses, being re-circulated through vents under the windows, see Figure 7. One can assume that the four windows that could be opened, on each bus, were open, but not widely, and did not let in too much air due to the cold temperature outside. Still, this outside air seems to have protected the passengers sitting next to the windows, see Figure 7, and this must be taken into account.

The concentration of small droplets/aerosols, containing the coronavirus, that the passengers in the bus are exposed to can be computed by using the ratio of the total volume of air in the bus to the volume of the cloud exhaled by the infected person, as in [10], see Appendix A. The useful reference concentration is that experienced by the person sitting next to the infected person in the bus, C15, C28 and C22 in Figure 7, after the infected person has exhaled. This concentration is clearly sufficient for these passengers to become infected over the span of those two 50 min. bus rides. The person sitting next to the open window in row seven and the persons sitting next to the aisle in rows eight and nine are presumably being protected by the (sinking) cold air coming through the window. The cloud exhaled by the infected person forms a bulb-shaped cylinder with radius shown in Figure 3. This cylinder is directed towards the center and rear of the bus (passenger C18), by the cold and warm airflow in row seven and aided towards the ceiling by the warm air flow in row nine, see Figure 8. The length of this cylinder is 2.75 meters, assuming that the width of the bus is 2.55 m. and the passengers are tightly packed with three rows measuring 2 m. and the space between the passengers being 0.75 m.

We know that the bus is close to 12 m. long and 3 m. wide, but unfortunately the exact dimensions of the bus are not available. Thus instead we use the dimensions of typical bus (Man Lion’s Coach) of this size, see https://www.dimensions.com/element/coach-buses, they are length 12 m., width 2.55 m. and height 3.81 m. Assuming that the height of the passenger compartment is 2/3 of the height of the bus, or 2.54 m., we get the volume 77.72 *m*^3^ for the interior of the bus.

Now the warm air vents under the windows are creating a flow across the bus, so the system length is the width of the bus, and we assume that the flow is the standard 0.25 m/s recommended for air-conditioning. This means that the Taylor Reynolds number is almost the same as that of the restaurant *Re*_λ_ = 705, so we can use the parameters computed in [10] for the restaurant. We extend the cylinder in the bus another 3 rows until it hits the ceiling and the air is sucked into the inlet of the air-conditioning, see Figure 8. Thus we get a cylinder of length close to 6 m. and radius 0.53 m. containing the droplets/aerosols exhaled by the infected passenger. This cylinder is mixed with the air from the rest of the bus and then blown out through the warm air vents since the air-conditioning is on re-circulation mode. Now the volume of the cylinder is 5.29 *m*^3^ and the total volume of the air in the bus is 77.72 *m*^3^ so the concentration gets decreased by a factor of 77.72/5.29 =14.69. Thus if the infected person has filled that cylinder once, by the time the air is blown through the warm vents, each passenger in the bus is exposed to a concentration of droplet/aerosols that is 1/14.69 of the exposure of the six passengers, sitting inside the bulb-like half of the cylinder, close to the infected person. However, the infected person fills the cylinder every 1.667 minutes and during the 50 min. ride, she has filled the cylinder 30 times. That means that the concentration that every passengers is exposed to is at the end of the ride 30/14.69 = 2.04 of what the people sitting next the infected person were exposed to initially, if no fresh air is entering the bus. At the end of the bus ride, it is almost as if everyone was sitting next to the infected person and gotten twice the infectious dosage.

However, fresh air is entering the bus through the four windows that can be opened and the persons sitting next to open windows seem to have been protected by the fresh air. Most of them did not get infected and of the two that did, their symptoms were mild, see Figure 7. If we assume that the airflow through the windows was enough to give half of the passengers the recommended fresh air, say 6 ACH, then the overall air exchange per hour (ACH) in the bus is 3 ACH and using the differential equation (A.3), in Appendix A, we get the concentration,

Aerosol concentration in the bus after 50 minutes = 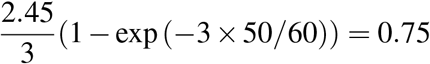, times the initial concentration (=1) next to the infected person, that each passenger is exposed to after 50 minutes. The fresh air cuts the concentration almost by a factor of three, but still leaves it high enough for most people to get infected. If everyone on the bus was getting the recommended dose of fresh air the concentration goes down to 0.42, and that might be low enough to avoid infection by many of the passengers, if the exposure time is not too long.

In [30] the outbreak on the bus is blamed on the re-circulating air-conditioning and the above analysis confirms that finding. One can look at the infection on the bus as a two-step process. First the infected person infects the passengers seating next to her or in the cloud she exhales and while it expands during the 1.2 second exhale event, in [10] this is called the vortex-building part of the cloud and forms a large bulb-shaped initial part of the cloud, see Figure 8. This part is highly infectious, may contain large droplets laden with the coronavirus, in addition to the small droplet/aerosols. It will infect anyone who is susceptible and exposed to it for a sufficiently long time. However, a limited number of people are infected and there is a delay time of 24-48 hours before they become infectious. We call this step one or the linear part of the contagion. The passengers are tightly crammed into the bus, with 0.75 m. between them, and the infected person infects 6 people (C15, C17,C18, C19,C22 and C26) exposed to her vortex-building cloud.

The second bus and the lay people at the temple that did not arrive by the two buses, form two control groups for the outbreak. Step one cannot occur on Bus 1, because no infectious person was on that bus, and they randomly scanned the 172 lay-Buddhists and 5 monks and did not find any infectious person there. Our infectious person on the other hand, did also effect step one infection at the ceremony, she was in close contact with at least seven persons, and probably sat next to two of them during a 30 min. meal, and exposed them to her vortex-building cloud. Thus step one can and did happen among the lay-Buddhists at the temple.

On Bus 2, there was also an infection involving many more people, that was not effected by close proximity (the vortex-building part of the cloud) and consists of the infectious person contaminating an environment, such as the interior of the bus (or the section of the restaurant, above). This a super-spreading event and we call it step two or the polynomial part of the contagion. In [30] the bus was split into two Zones 1 and 2, see Figure 7. It was argued that the passengers in Zone 2 were outside the contagious vicinity of the infected passenger and had to be infected by airborne droplet/aerosols. Indeed, based on the above analysis, we would argue that the passengers C15, C17,C18, C19,C22 and C26, see Figure 8, were infected by a step one contagion and everyone else infected on the bus was infected by a step two contagion. They were infected by aerosols, since the small droplets quickly evaporate leaving aerosols, carried by the air being re-circulated through the warm vents, by the air-conditioning on the bus. The air-conditioning thus was the super-spreader in the confined space of the bus interior. Step two cannot happen on Bus 1 because there is no infected person. Step two cannot happen at the temple because the ceremony is outside and the lunch is too short in duration and the lunch hall too spacious for the concentration to build-up.

Our study has several limitations that should be pointed out. First, we do not have the precise dimensions of the bus and the standard bus is designed to carry 50 persons. However, we know that the bus was of roughly the standard size and the passengers tightly crammed in with 0.75 m. between them and three rows extending to 2 m. Thus our guess is probably close. The difference that the correct dimensions would make is that the Taylor-Reynolds number and the energy dissipation, that determines the time scale, would change slightly. The effect would be small. Second, if all the small droplets are heated by the air-conditioner and blown out through the warm vents they would evaporate leaving behind droplet nuclei or aerosols only. However, this is probably an oversimplification, a portion of the contaminated cloud sinks at the back of the bus and re-circulates along the floor to be mixed-up into what is called in [10] the vortexstreching part of the cloud. Thus a small portion of the small droplets may continue to survive and form a part of the atmosphere in the bus. It was shown above that injecting the recommended air for all the passengers on the bus reduced the concentration to 0.42 times what a passenger sitting next to the infected person would experience. Putting in twice the recommended amount of air reduces it to 0.21 and slightly more than tripling the recommended amount of air, by opening all the windows wide, reduces the concentration to 0.14 times what it is, sitting next to the infected person. This may explain why no infections have been registered the New York subway, see https://www.nytimes.com/interactive/2020/08/10/nyregion/nyc-subway-coronavirus.html and https://www.nytimes.com/2020/08/03/nyregion/nyc-subway-coronavirus.html, where the airflow is three times the recommended amount. Most modern buses take fresh air through the ceiling rather than the windows. This is probably also the case for the bus in our study but it does not change the analysis. The cold air still falls to the floor, and protects some of the passengers on the way, whereas the warm air rises and presumably the ventilation system did not let in too much cold air because of the temperature outside. The example of the Norwegian bus, see https://www.fhi.no/en/news/2020/new-coronavirus-variant-contributed-to-outbreak-among-coach-trip-passengers/, shows that even low concentration can infect everyone on a bus if the exposure time is long enough.

## 4 The Call Center in Seoul

On March 8th 2020, the Seoul Metropolitan Government was called to a Call Center in Seoul Korea, where an outbreak of Covid-19 had occurred. On March 9, the Korean CDC and local governments formed a joint response team and launched a epidemiological investigation with contact tracing. The outbreak had occurred on the 11th floor of a 19 story building, the first 11 of which contained commercial offices and the top seven residential apartments. The Call Center occupied four floors, 7th through 9th and the 11th floor. It employed 811 people in total, 216 of whom worked on the 11th floor. There was no regular contact between the people working in different floors of the Call Center. The workers however met casually in the elevators and in the lobby of the building. The investigation revealed a single person on the 10th floor developing symptoms on February 22nd, see Figure 9, the source of this person’s infection is unknown; on Febuary 25th a single person on the 11th floor developed symptoms, he or she had presumably been infected by contact with the person on the 10th floor, and three days later, on February 28th, three more people on the 11th floor developed symptoms. On February 29th and on March 12th two more people on the 10th floor and on March 2nd one person on the 9th floor developed symptoms. These infections where presumably caused by contact with people on the 11th floor, but they did not spread further on the 9th and 10th floor and will not be discussed further below.

All of the infections that we discussed so far have been step one contagion, using the criterion of the analysis of the buses above. They were caused by a close contact with an infected person over a sufficiently long time. However, on March 4th a sharp increase in the number of infections starts and persists for six days, see Figure 9, this increase is multiplicative, not just additive as the cases discussed above, and seems to signal the onset of a step two, polynomial, contagion. The other noteworthy thing about the outbreak is that is seems to occur mostly on one side of the building, see Figure 10, as noted in [24]. Of the 94 people infected on the 11th floor, 89 work in a big room, Room 1, on the top of the diagram in Figure 10 (presumably there is a missing row with 10 infected people in Room 1) and 4 work in a smaller room, Room 2, at the bottom. One works in a separate office space at the bottom right corner, see Figure 10. Moreover, all the infected persons in the smaller room are isolated, they do not have other infected people around them. Thus it looks like step one and step two contagions took place in Room 1, but only a step one contagion took place between Room 1 and Room 2, and only a limited (no neighbors infected) step one contagion took place in Room 2. Notice that step one contagion involves a limited number of individuals and has a time delay of 1-2 days, for example Feb. 25 to March 2nd on Figure 9, but step two contagion is much faster and involves a large number of individuals, see March 4 to March 9 on Figure 9, it also ends on March 9th when the building is closed. We will now use the air-flow analysis to try to figure out what the explanation of this could be.

The information that we have obtained about the dimensions of the building, where the Call Center is located, and the dimensions of Rooms 1 and 2 are not exact but close estimates, so we can make an educated guess that permits a qualitative and approximate, if not quantitative, estimate. The dimensions of the building are approximately 23 by 23 meters and Room 1 approximately is 23 meters long and 7 meters wide with 2.25 height to the ceiling, Room 2 is approximately 16 meter long and 6 meter wide also with a 2.25 meter ceiling height. This give the volume 362.25 *m*^3^ of Room 1 and 216 *m*^3^ of Room 2. The air-conditioners cause the air to blow across the room, we assume the standard recommended (Lagrangian) wind is 0.25 m/s. The vents blowing the air into the room are located in the ceiling and presumably close the the center. Thus the system length is half the width of the room and this makes the Taylor-Reynolds number *Re*_λ_ = 705 for the small room and a little larger for the big room. We will use *Re*_λ_ = 705 for both rooms. This allows us to use the same parameters as for the restaurant and the bus. Now the extent of the exhaled cloud is the same 0.25 m., so an infected person has to breathe 7/0.25 = 28 times to fill a cylinder extending 7 meters across Room 12. The breathing happens every 5 seconds, so the time required is 5 times 28 = 140 second or two and a third minutes. In one hour the infected person has filled 60/2.33 = 25.75 cylinders. The volume of the cylinder with the droplet/aerosols containing the virus is (0.53)^2^ ×π ×7 = 6.18 *m*^3^. This gives the concentration 6.18/362.25 = 1/58.63 in two and one third minutes, as a fraction of the concentration sitting next to the infected person, in Room 1. In one hour this builds up to be the concentration 25.75/58.63 = 0.44. After an 7 hour workday it is 3.1. The computations for Room 2 are similar. In two and one third minutes we get 6.18/= 1/34.95 and in one hour the concentration 25.75/34.95 = 0.77. After 7 hours the concentration in Room 2 has built up to 5.1 of what it would be sitting initially next to the infected person.

The situation is much better with the recommended air-conditioning, that was present in the Call Center at the time of the outbreak. We use the differential equation (A.3), in Appendix A, to compute the concentrations assuming the recommended 6 ACH (air exchanges per hour). Since it was cold outside we assume that 50% outside air was entered into the system and the filtration was not of high enough quality to filter the aerosols, so they were recirculated into the rooms. The concentration in Room 1 becomes 0.1 after one hour and 0.15 at the end of the workday. In Room 2 it is 0.2 after one hour and 0.26 at the end of the workday. We can now construct a reasonable history of the outbreak. The person who became symptomatic on February 25 infected 3 others sitting next to him/her by step one contagion, the step one contagion spread to the adjacent tables where these persons were sitting and several days before March 3rd more than three persons had been infectious simultaneously. This gives the concentration *>* 3 × 0.15 = 0.45, that is contagious at end of each of these workdays. In fact it is already 0.3 after one hour. The result is step two contagion on March 3 that grows polynomially in Room 1 and infects half of the workers there. Now why does this not happen in Room 2? The most likely explanation is that the contagion never reached a critical mass in Room 2. The workers in Room 2 probably got infected by casual contact with the workers in Room 1. Once a large number had been infected there, by a step one contagion, they did not have time to build up the concentration in Room 2, before the building was closed, to see a step two contagion. Thus the concentration stayed low in Room 2 and even the step one contagion did not spread there.

The analysis above shows that the contagion in Room 1 in the Call Center, spread initially by personal contact or a step one contagion, but once the number of infected people in Room 1 hit a critical mass, with three or more people infectious, it seeded a step two contagion. The infection then quickly spread to the whole room by the air-flow in the room driven by the ventilation. The step two contagion stopped on March 9th, when the building was closed but people continued to be develop symptoms for another 11 days thereafter, some of whom may have been infected by personal contact or step one contagion.

The initial explanation, see [24], was different: It was thought that everyone in Room 1 got infected by casual contact, or step one contagion. The problem with that explanation is that it would simply have taken too long a time to spread to all the tables in Room 1. Figure 9 makes this clear. From February 22nd to March 3rd a period of 10 days, 10 onsets of symptoms took place, presumably all by individual contact, see Figure 9. After March 3rd a cascading infection took place that can only be explained by a step two contagion. It resulted in the onset of symptoms for 76 persons and the whole event took place in six days, see Figure 9. After the building was closed on March 9th the rapid rate of infection stops abruptly: Only 11 onsets of symptoms occurred during the subsequent period of 11 days, see Figure 9. Some if these had already been infected but some may have been infected by a much slower step one contagion, caused by individual contacts outside the Call Center. Thus two step one contagions of 21 people took 21 days whereas one step two contagions of 76 people took only 6 days. If the step two contagion had happened at the same rate as the step one contagion, or roughly one person per day, it would have taken 76 days, or more than two months to infect all the people that it infected. Instead it took only a week.

## 5 Conclusions

The analysis of the three outbreaks of the SARS-CoV-2 Coronavirus infections discussed above shows that all of them involved a close contact infection, step one, and a subsequent step two infection that happened in spite of physical distancing and thus must have been effected by small droplets and aerosols. The reason for the size of the outbreak in all three cases seem to be the build-up of the droplet/aerosols carrying the SARS-CoV-2 Coronavirus in a confined space over time. In fact, the small droplets evaporate quickly leaving only aerosols. This has implications for small and large spaces where ventilation may not be sufficiently effective to remove the virus-containing aerosols.

The analysis summarized above make it clear that the part of the restaurant in Guangzhou where the outbreak took place was extremely vulnerable to transmission by aerosols. The aerosols were not being distributed to the whole space in the back of the restaurant (the well-mixed hypothesis did not apply). The air flow from the air-conditioner was restricting the space that needed to be ventilated, to the pyramid in Figure 5. The vents were closed so no fresh air was entering this space and the air-conditioner did not have filters capable of filtering out the aerosols. Gentle heating probably accelerated the evaporation of small drops into aerosols. These factors conspired to make the ventilation coefficient *k*_*v*_ = 0 in Equation A.3. The settling coefficient of the aerosols, that were being blown around and re-injected into the pyramid by the air-conditioner, was also *k*_*s*_ = 0. The only mitigating influence was the decay coefficient of the virions in the aerosols, that is very small *k*_*d*_ << 1, see [10]. This decay coefficient was made smaller by the low relative humidity in the restaurant. The combination of these factors made the back of the restaurant in Guangzhou a very infectious place in a short time. In addition, the density of patrons was high due to the Chinese New Year celebration.

The environment inside the bus in Ningbo city is in many ways similar to the back of the restaurant in Guangzhou. The air-conditioning is in re-circulation mode, it does not let in any outside air and does not have filters of sufficiently high quality to filter the virus-carrying aerosols. The dimensions of the bus are such that the Reynolds number of the Lagrangian flow is similar to that of the restaurant and the density of passengers is also very high due to the extra seat in very row. Thus the inside of the bus traps the aerosols and their concentration increases during the bus ride. However, there is a difference due to the ventilation provided by the open windows on the bus, that that is not present in the restaurant. This difference shows how important ventilation is and what an effective tools it can be to prevent contagion. In Figure 11, we compare the aerosol concentration for the three cases studied in this paper. The attack rate in Bus 2 (35 %) is similar to that of the restaurant (43 %), but this is due to the exposure time that is twice as long (two times 50 minutes) in the bus. Since the concentration is considerably higher in the restaurant, it may indicate that after a threshold concentration is reached, the exposure time is strongly correlated to the infection rate.

The Call Center in Seoul is different from the previous two cases because it has greater ventilation. However, because the infection reaches critical mass with more than three persons simultaneously infected the ventilation is overwhelmed. The density of people in the Call Center is also very high, the ventilation is not strong enough to service the number of people in the Center nor to prevent the outbreak of infections. The long exposure time also plays a role, although the concentration is smaller than in the previous cases, see Figure 11, it is still enough to cause most people to become infected. The attack rate on the 11th floor in the Call Center was 43.5 %, but in Room 1 it was considerably higher at 59.3 %. It is likely that if the Center had not been closed, when it was, everyone in Room 1 would have been infected and a step two outbreak would also have occurred in Room 2.

These three cases have in common that the outbreak involving a large number of infections is caused by aerosols, whose concentration builds up at a polynomial (in time) rate. This it is due to lack of proper ventilation, insufficient filtration and re-circulation of the aerosols into the contaminated space. The amount of ventilation varies, from being absent in case of the restaurant, to being considerable in the case of the Call Center. This is brought out by the dramatic differences in the build-up of the concentrations in Figure 11. But the beneficial influence of the ventilation is countered by an increase in exposure time, in case of the Bus and the Call Center, and the presence of a critical mass of infectious person in the Call Center. In all three cases, the high density of people in these confined spaces also plays a role.

It is also clear that a sufficiently strong ventilation could have prevented the second step contagion for the persons who were sufficiently far away from the infected person. On the bus it might have been enough to open the windows wide to triple the airflow. In the Call Center tripling the recommended fresh air-flow would have cut the concentration down to 0.15 after an eight hour workday, with three infectious persons present. This is probably enough to prevent most infections. Only in the restaurant does this seem difficult, since 5 times the amount of fresh air is needed to decrease the concentration to non-infectious levels. But an installation of an additional air-conditioner pushing air, perpendicular to the air-flow in the space where the contagion took place would have changed the situation. Then the whole air-space of the restaurant would have been used (the well-mixed hypothesis would apply), as in the Call Center, and the tripling the recommended amount of fresh air in the whole restaurant might have sufficed to prevent the outbreak.

Finally, we can redo the analysis assuming that everyone in the confined space is wearing a mask. The difference between the amount of drops and aerosols, with and without a mask, is studied numerically in [14]. The study concentrates on coughing with zero ambient fluid velocity but can still be applied to our study, with some assumptions. The striking effect is how much the volume of the large drops is reduced by a quality mask. The droplet volume can be reduced by a factor of 6-7, see [14]. This has important implications for the step one contagion discussed above. If both the infected person and everyone in his or her vicinity wears a mask, the concentration of drops can be reduced from 1 to 0.15. This is probably enough to prevent infections if the exposure time is small. The effect of a mask on the distribution of the small droplets and aerosols is smaller, see [14]. These small droplets and aerosols can still get around the mask but in a reduced volume. We will make the assumption here that their volume is reduced by a factor of 3. This assumption is consistent with the results in [14] but needs more experimental verification. If everyone in the restaurant had been wearing a mask and the air-conditioning had followed recommendations, the concentration of the aerosols would have been 1.57, after one hour, for everyone sitting at the three tables A, B and C. The concentration is reduced to 0.52 by the mask, that person inhaling the droplets and aerosols is wearing. This is probably still high enough to be contagious. On the bus the concentration of the droplet/aerosols would be reduced to 0.25 after 50 minutes. The concentration will be further reduced to 0.08 when inhaled through a mask. This is probably enough of a reduction to prevent the step two contagion. In the Call Center with three infectious persons present the concentration is reduced to 0.15 (with three infectious persons present) after an 7 hour workday, in Room 1, and to 0.05 when inhaled through a mask. This may have sufficed to prevent the (step two) outbreak in the Call Center. In Room 2 it becomes, 0.09 after a 7 hour workday and 0.03 when inhaled through a mask.

The silver lining of our study is that if physical distancing is practiced, everyone wears a quality mask and the ventilation in confined spaces is sufficiently improved, the outbreaks of the types discussed above can be avoided, and perhaps most infections by the SARS-CoV-2 Coronavirus can also be avoided. The problem is that even if people wear a mask and follow the distancing rules, they may enter confined spaces where the concentration of aerosols carrying the Coronavirus may have reach infectious levels, as in the three cases studied above.

In [25] the transmission of the virus droplet/aerosol through an air-conditioning system is discussed with the conclusion that greater volume of outdoor air and MERV-13 or HEPA filters, with the capacity of filtering out the droplet/aerosols, need to be used. This is consistent with our observations. However, the air-conditioners in current use in most buildings may be unable to use MERV-13 filters or have fans that are not able to handle the required volume of outdoor air. Indeed a new generation of air-conditioners that meet these requirements may be needed.

## Data Availability

All data used is from published sources.

## Acknowledgements

The author wants to thank Klaus Schauser and Albert Oaten for enthusiastic support, Inga Dóra Björnsdóttir for careful editing and Ken Beisser for expert advice on air-conditioning. We also thank Knut Bauer for providing us with the graphic illustration in Figure 6 and the authors of [30] for making their results available. He especially grateful to Luiza Angheluta for reading the manuscript and making useful comments and to Youngdon Lee for making a video of the Call Center in Seoul.

## A Method

We use the Richardson distribution of passive scalars to understand the shape and extent of the cloud of small droplets and aerosols, exhaled by a person infected by the SARS-CoV-2 Coronavirus, see Appendix A, in [10]. The theory of passive scalars and the leading form of the Richardson distributions have been known for some time, see see [21] and [3]. It is

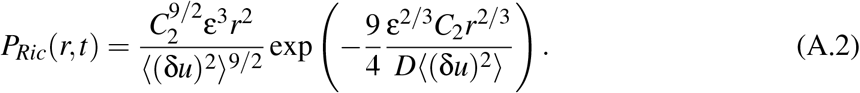

*r* is the distance between the particles (droplet/aerosols), *C*_2_ is the coefficient of the second structure function of turbulence, ε is the energy dissipation, δ*u* is the velocity difference (in the structure functions), and *D* is the coefficient in the Richardson law, see [10], Appendix A. For *r* large, the additional terms in the Richardson distribution are small, see [28].

There are two problems with the formula (A.2) that make it less explicit than it seems: First, it requires a formula for the average of velocity differences ⟨ (δ*u*)^2^⟩, that is the second structure function of turbulence, as a function of Lagrangian time. Second, one must be able to evaluate the coefficient in the exponential part of (A.2). The numerical form of ⟨ (δ*u*)^2^⟩ has also been available for some time, see [28], but in [2] the author and his collaborator were able to develop a theoretical model inspired by [8] and [17]. We employ the theoretical structure function *S*_2_ from [2] and use the stochastic closure theory (SCT), see [6, 7, 9], to compute the coefficient *C*_2_ in the exponential of the formula for the Richardson distribution (A.2). This computation, that interpolates the coefficients computed in [20] for the Reynolds number *Re*_λ_ = 705, is explained, in Appendix B, in [10]. This value of *Re*_λ_ gives the coefficient in the Richardson distribution 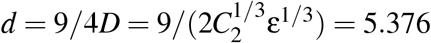

The conventional way of modeling the concentration C, of droplets and aerosols in a room, is by a differential equation

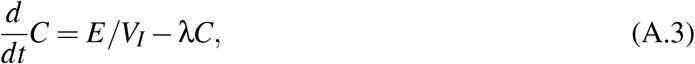

where E is the exhalation rate, in dimensions droplets and aerosols per volume per minute, and *V*_*I*_ is the volume of infectious air. In the example of the restaurant above *V*_*I*_ is the volume of the pyramid and not the volume of the whole airspace. This means that the frequently made hypothesis, that the whole available air space is well mixed, does not hold for the restaurant and may not be true in other cases. λ is the decay coefficient of the concentrations, it is composed of a sum of three decay rates λ = *k*_*v*_ + *k*_*s*_ + *k*_*d*_, where *k*_*v*_ = 1*/t*_*v*_, *k*_*s*_ = 1*/t*_*s*_, and *k*_*d*_ = 1*/t*_*d*_. *t*_*v*_ is the time it take to ventilate the room, *t*_*s*_ is the settling time of the droplet/aerosols and *t*_*d*_ the decay time of the viruses in the aerosols.

The results of the simulations for the restaurant in Guangzhou are shown in Figures 2-3. In Figure 2 we show the Richardson distribution as a function of time and space. The distribution is plotted in unitless temporal and spatial variables. But each temporal unit is τ_η_ = 3.55 ms (milli-seconds) and the exhale event takes 1.2 seconds = 3.55 × 340 unit steps. The time scale is computed from the dissipation rate ε by the formula τ_ε_ = (ν3*/*ε)^1*/*4^, where ν is the viscosity in air at 20 C^*o*^. Figure 3 (left) is a cross-section of the exhaled cloud of droplet/aerosols from the center to the boundary. The blue part is the emitted vortex-building part the cloud, the Lagrangian eddies spin up to the maximum radius (particle separation) of the cloud and then begin their slow decay in the Richardson cascade. However, at the maximum a faster process of vortex stretching associated with a different scaling, see [2], begins and the radius of the cloud shrinks and the vortex-streching eddies (the red part of the cloud) build, reach their maximum and then shrink by the vortex-streching, their spinning also speeding up. We call the first part of the cloud the vortex-building (extensive) part of the cloud and the second part the vortex-streching (less extensive) part. Figure 3 (left) shows the radial extent (y-coordinate) of the cloud as it is created by the velocity of the exhaled breath (1.5 m/s). This assumes that the vortices are created by the velocity of the exhaled air, but then evolve by the Lagrangian evolution, see [10] for more details on this assumption. To get the spatial extent (x-coordinate) of the cloud we have to compute how far the particles are translated in a given time interval. This is determine by the Lagrangian velocity of the cloud, that is 0.25 m/s, or in 1.2 seconds the particles are translated 1.2 × 0.25 = 0.3 meters. This means that the length of the cloud in Figure 3 (left) is 2 × 0.3 = 0.6 meters. On the right hand of Figure 3, we show how the concentration, inside the pyramid in Figure 5, builds up over time. Notice that one, the normalized level at the infected person, is reached after 12 minutes. After one hour the concentration has reach more than five times that level.

**Figure 2:**
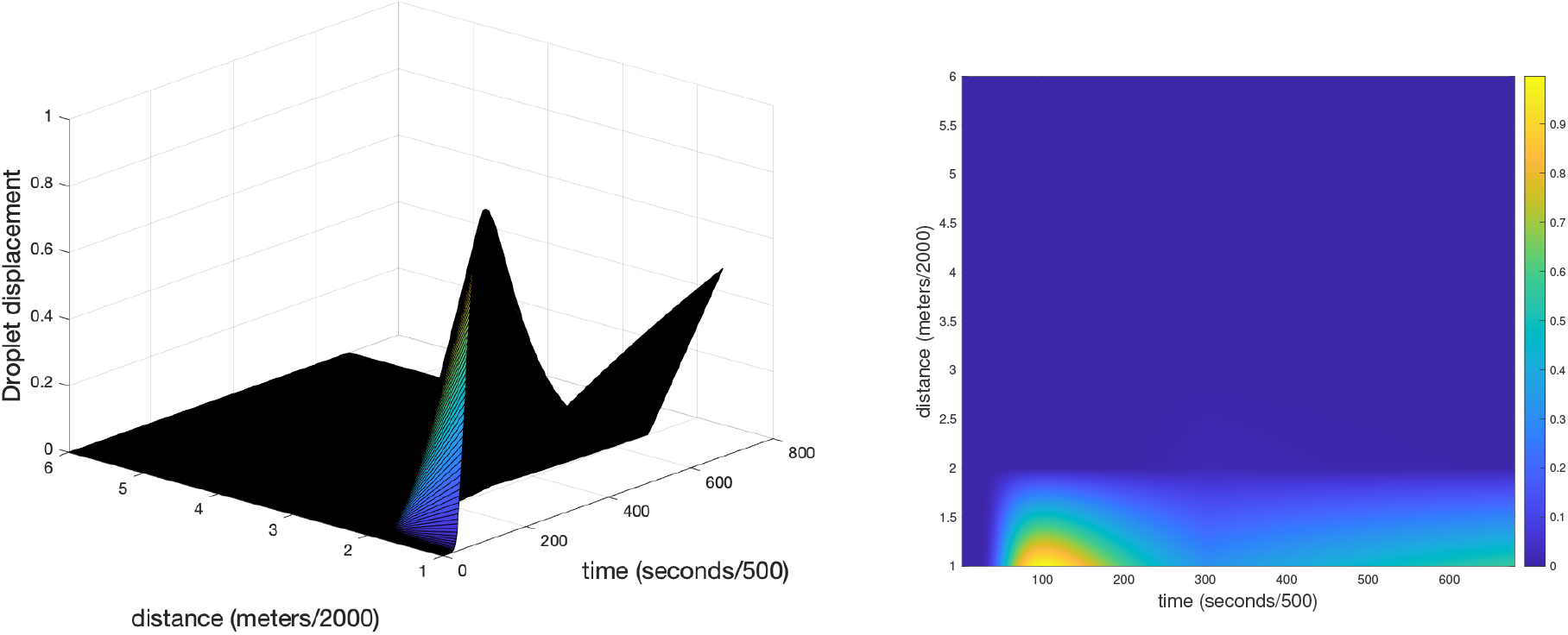
The Richardson distribution for the particle separation. The Figure is taken from [10].

**Figure 3:**
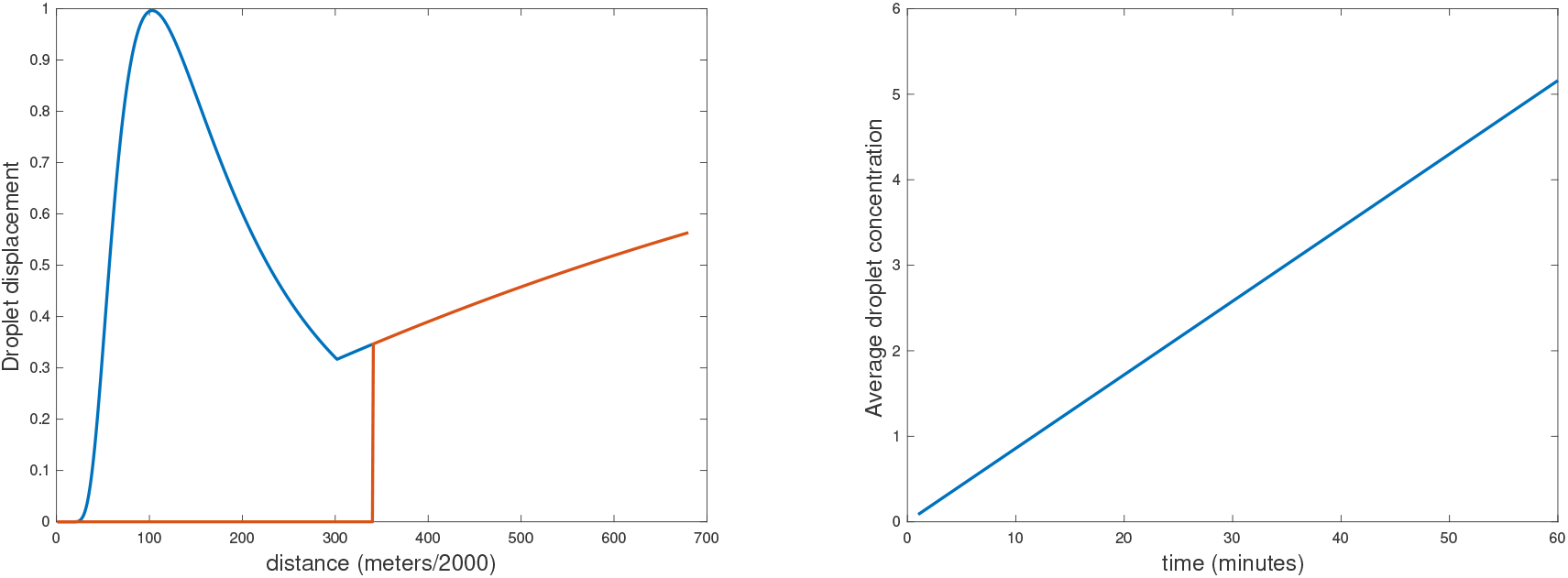
(Left) The total distribution of droplet displacement after 2.4 second of exhaling. The blue (vortex-building) part corresponds to the first 1.2 second of the exhale event and the red (vortex-streching) part the subsequent 1.2 seconds. This is a cross-cut giving the longitudinal shape of the, cylindrically symmetric, exhaled cloud. (Right) The average, in space (inside the pyramid), droplet concentration as a function of time, in minutes, normalized by the concentration at the infected person. The concentration builds up to one (the concentration at the infected person) in 12 minutes and 5.16 times that in an hour. The Figure is taken from [10].

**Figure 4:**
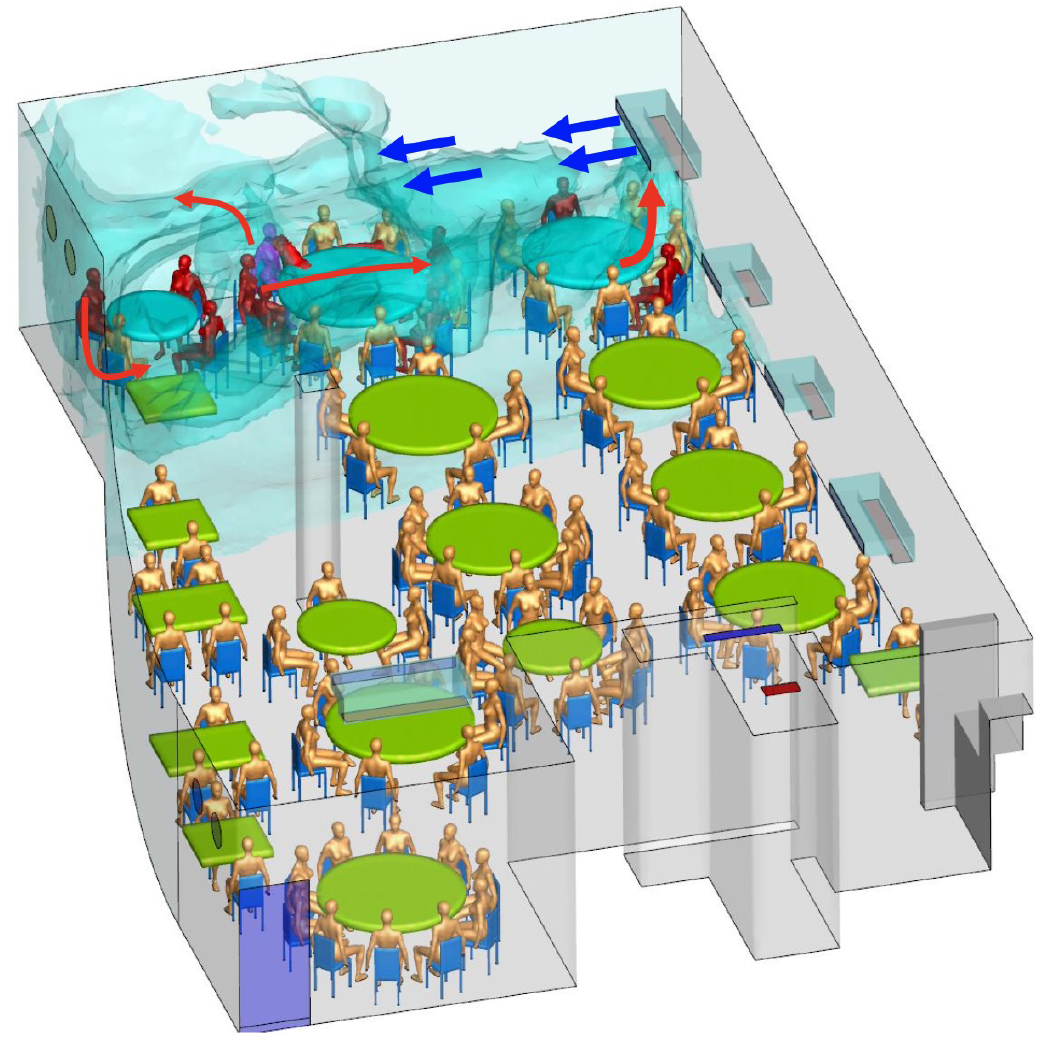
A CFD simulation of the contagion in the restaurant in Guangzhou China. The blue gas models the contamination by the droplet/aerosols. The infected persons are colored red. Notice that the contaminated region roughly forms a pyramid with base on the wall opposite to the air-conditioner. The figure is taken from [22].

**Figure 5:**
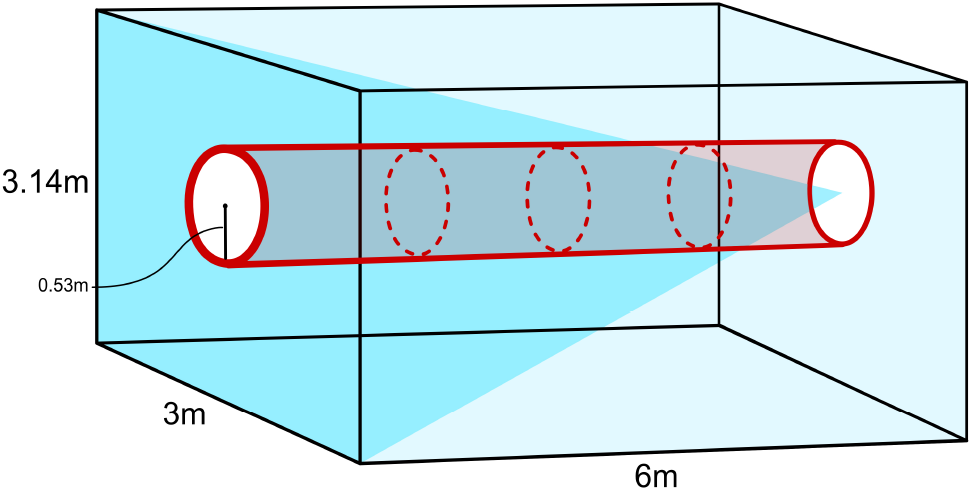
A cartoon (not to scale) of the volume (red cylinder) where the droplet/aerosol clouds propagate and the volume (blue pyramid) that the droplet/aerosol clouds get spread to by the air-conditioning. This figure is taken from [10].

**Figure 6:**
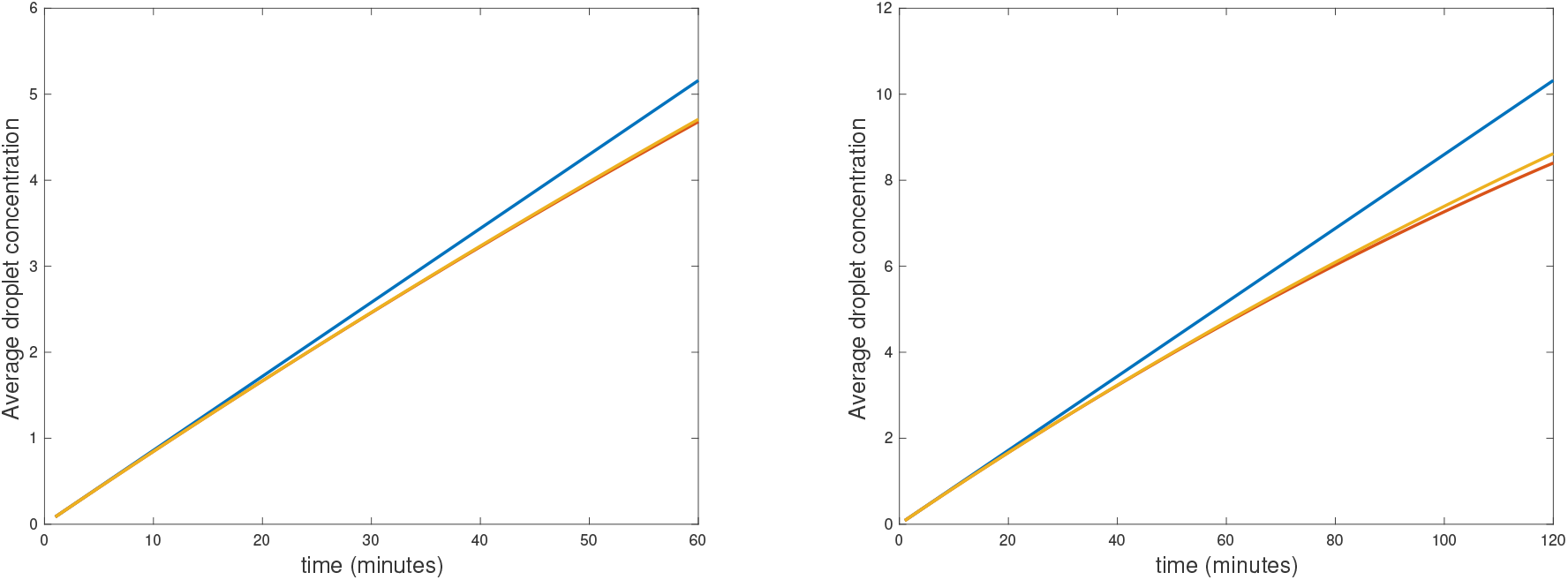
(Left) The aerosol concentration for *one hour*, top (blue) linear curve, middle (orange) solution of the ODE, bottom (red) the quadratic polynomial. The last two lines are hardly distinguishable. (Right) The aerosol concentration for *two hours*, top (blue) linear curve, middle (orange) solution of the ODE, bottom (red) the quadratic polynomial. The last two lines begin to separate on the right. This figure is taken from [10].

**Figure 7:**
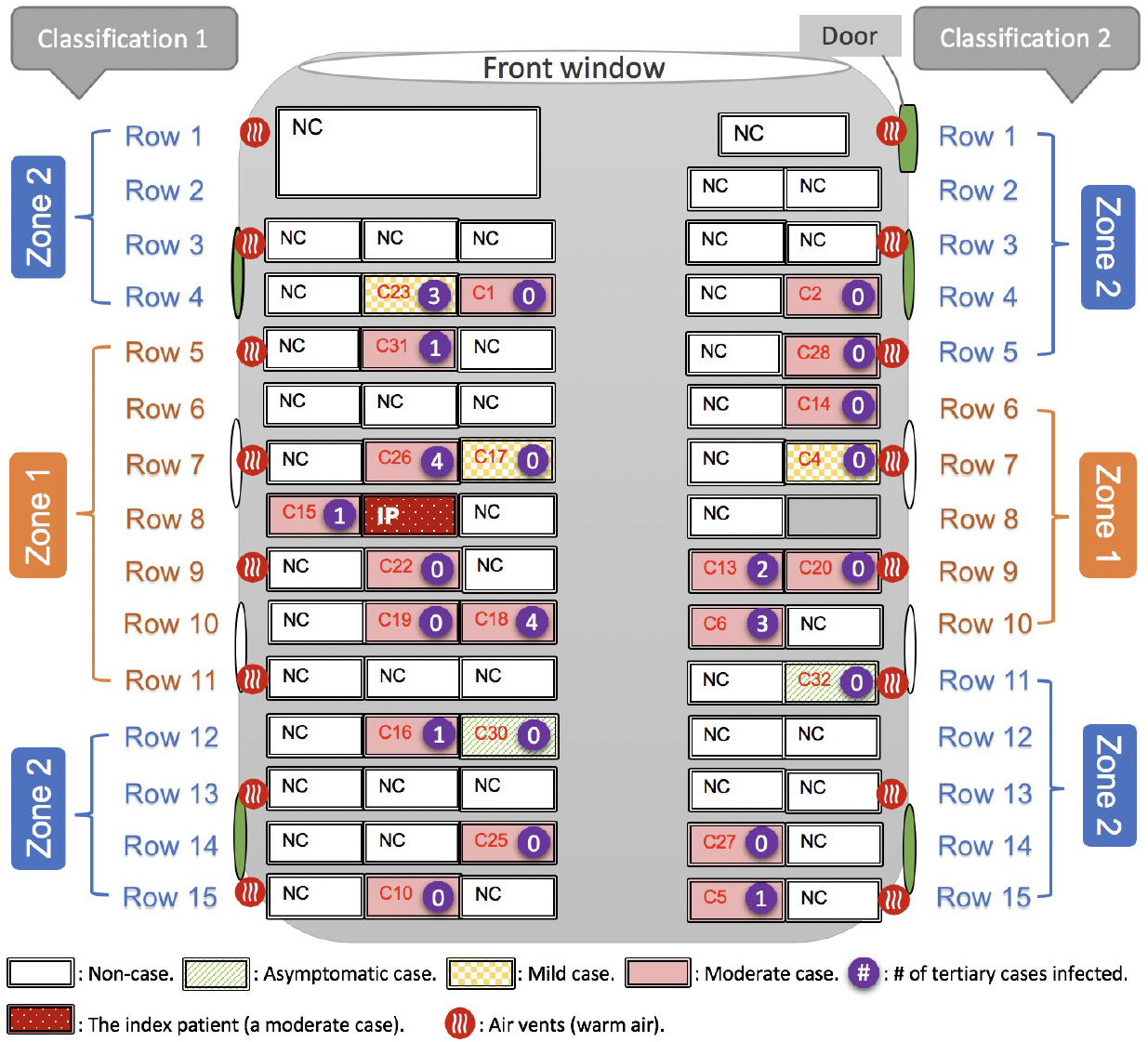
A schematic diagram of the bus indicating where the contagious person sat (red) and where the infected paseangers sat (pink, green and yellow). The remaining passengers (white) and the bus driver were not infected. Notice the warm-air vents under the widows (red with white squiggles) and the openable windows colored white, the windows that cannot be opened are colored green.The zones indicate distance from the infected person. The figure is taken from [30].

**Figure 8:**
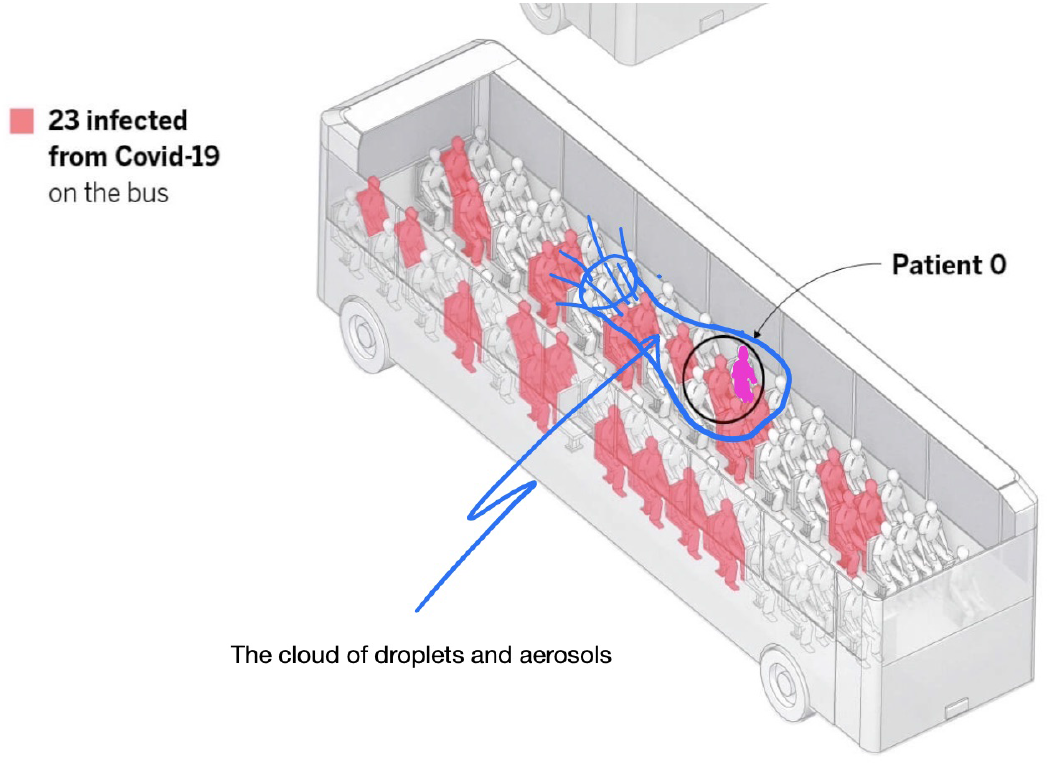
The cloud of droplets and aerosols emanating from the infected person on the bus. The infectious person is circled and marked as patient 0. The passengers that got infected are colored pink. The figure is taken from El País and modified.

**Figure 9:**
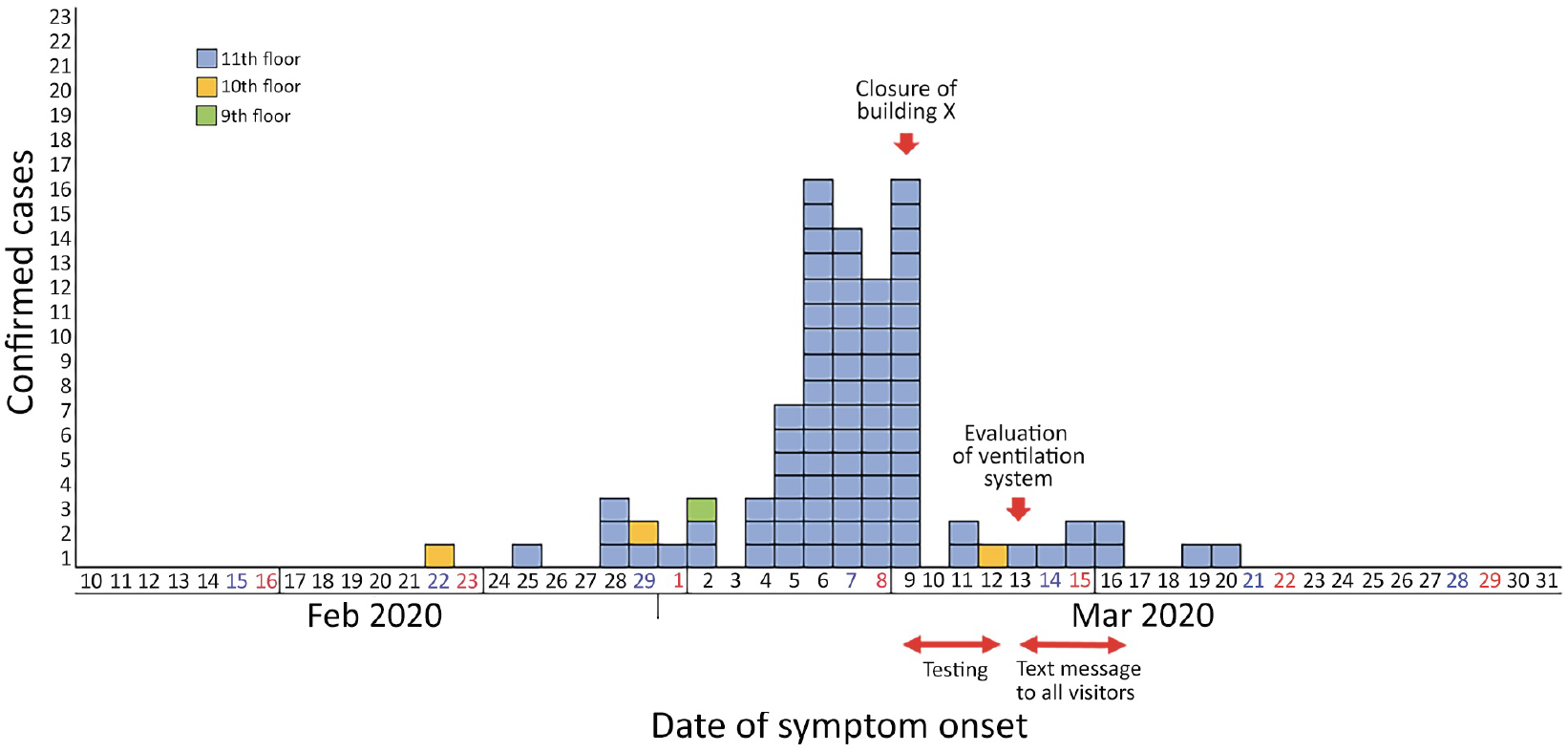
The number of coronavirus infections by date in the Call Center in Seoul, Korea 2020. Yellow denotes 10th floor, blue the 11th floor and green the 9th floor. Notice the polynomial increase starting on March 4th. The figure is taken from [24].

**Figure 10:**
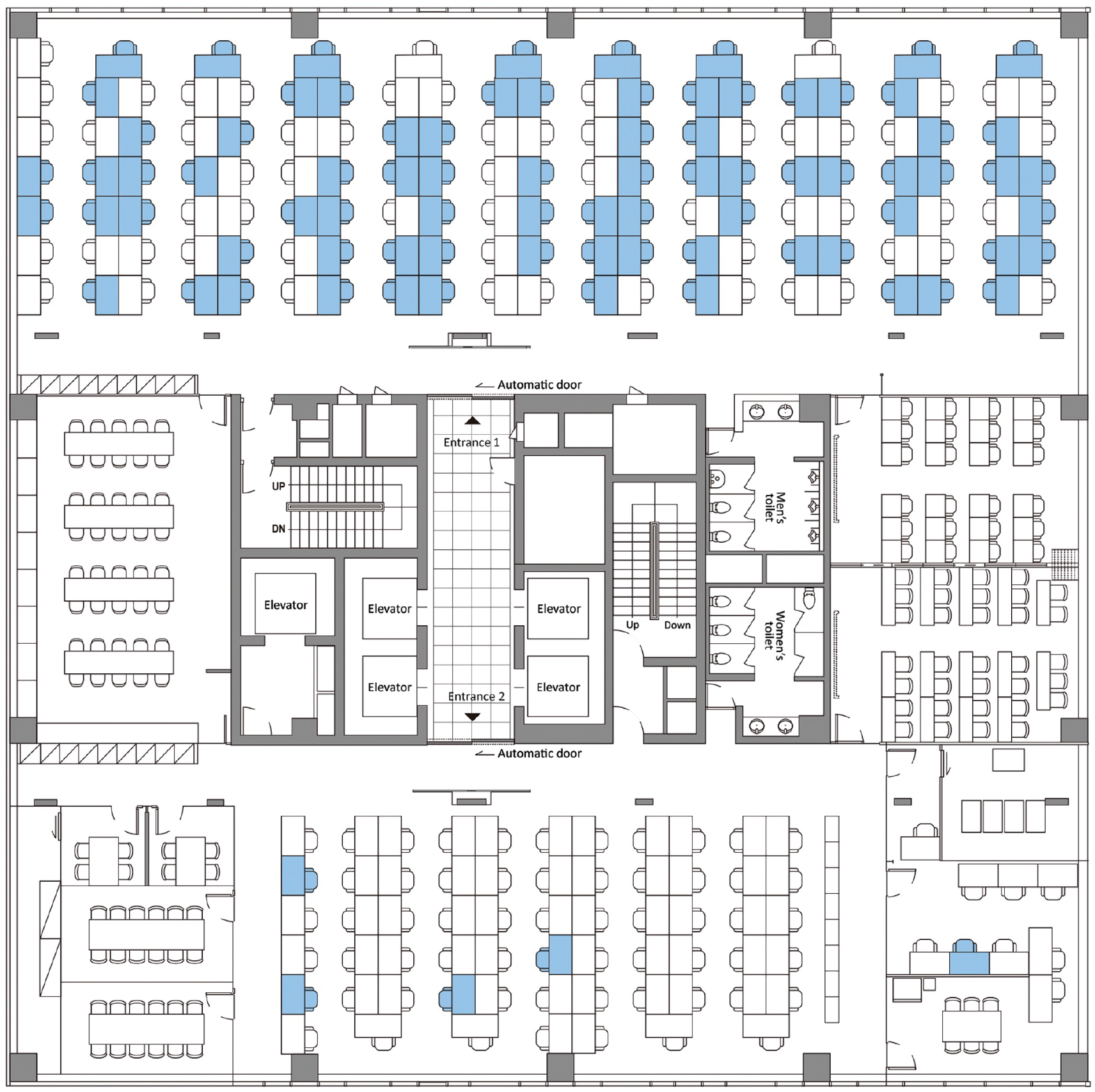
A schematic diagram of the Call Center indicating where the infected persons sat (blue). One row of of 13 seats with 10 infected people is missing at the top.The figure is taken from [24].

**Figure 11:**
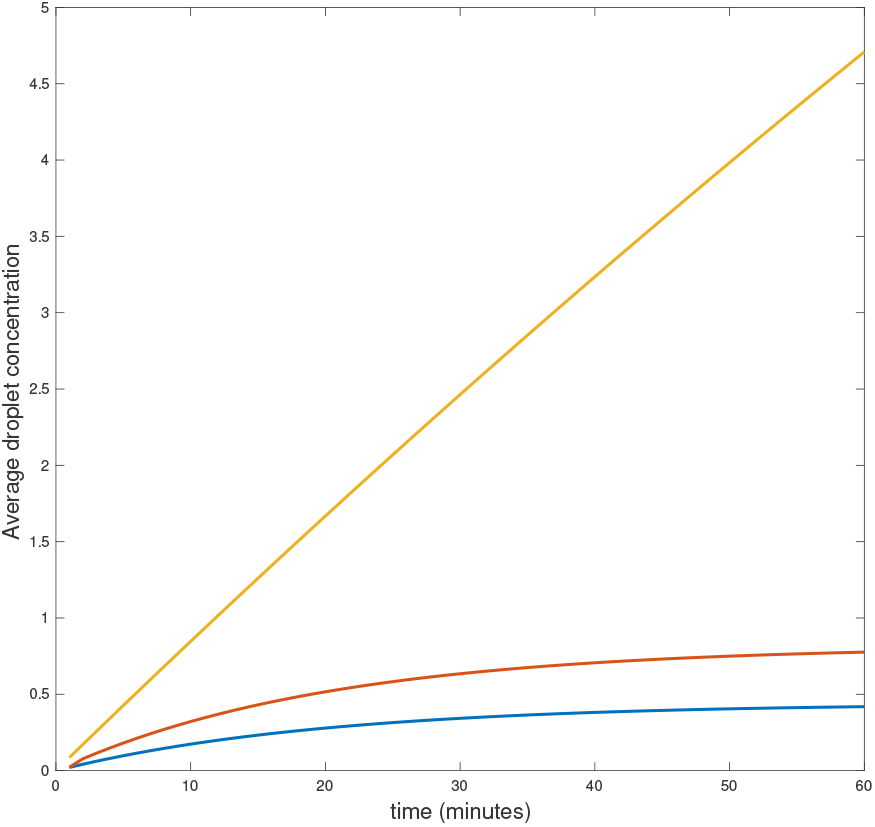
Aerosol distribution as a function of time in minutes. Top: restaurant, middle: bus, bottom: Room 1 in Call Center. The figure illustrates how infectious lack of ventilation is, compared to some ventilation, but also how lower concentrations can still build to infectious levels over time.

